# A Randomized Controlled Trial Evaluating the Effect of Adalimumab Upon Pre-Clinical Biomarkers of Cardiovascular Disease in ACPA-Positive Rheumatoid Arthritis

**DOI:** 10.1101/2025.01.26.25321158

**Authors:** Stephen P Oakley, Alison J Gibberd, Christopher Oldmeadow, Ranjeny Thomas, Hanno Nel, Niloofar Esmaili, Lyanne Weston

**Author notes:** **Corresponding author:** Dr Stephen Oakley, Staff Specialist Rheumatologist, John Hunter Hospital, Lookout Road, New Lambton, NSW 2305, AUSTRALIA.

## Abstract

**Objectives:** Rheumatoid arthritis (RA) is associated with elevated risk of cardiovascular (CV) events that has been attributed to inflammation although this is not well supported by randomised controlled trial (RCT) data. This RCT evaluated the effect of adalimumab upon endothelial function and arterial stiffness in ACPA-positive RA.

**Methods:** Sixty subjects with active ACPA-positive RA stratified as Early (<12 months n=30) or Established (>12 months n=30) were enrolled and randomised 1:1 within each group to receive 40 mg adalimumab or placebo. They were assessed by DAS28-CRP, Reactive Hyperaemic Index (RHI) and Carotid-Femoral Pulse Wave Velocity (CF-PWV) twice prior to treatment and then at 1, 4, 12 and 24 weeks with one final open-label at 36 weeks. Secondary analysis evaluated the effects of disease duration, Shared Epitope (SE) and smoking status.

**Results:** There were significant treatment effects upon DAS28-CRP at weeks 1, 4 and 12 in Early RA with transient effects upon RHI at week 1 in Established RA and CF-PWV in Early RA at week 4. Area-under-the-curve analysis found positive treatment effects in subjects expressing one SE (DAS28-CRP p 0.031, CF-PWV p 0.034) and in non-smokers (DAS28-CRP p 0.096, CF-PWV p 0.033).

**Conclusions:** This RCT appeared to show positive treatment effects upon CV risk. However, the effect of adalimumab upon RHI may be type 1 statistical error and the effect upon CF-PWV more likely represents physiologic pain-driven mechanisms rather than structural effects. This study highlights the challenges, the limitations, strengths of and opportunities for CV biomarker research in this complex field.

## Introduction

Rheumatoid Arthritis (RA) is a chronic destructive autoimmune inflammatory arthritis that is associated with 50% increase in cardiovascular (CV) morbidity and mortality^1^ primarily in “seropositive” RA^2^. Within RA populations greater inflammatory burden is associated with greater risk of CV events^3^ and more severe pre-clinical CV disease (CVD)^4^. Higher CV risk is also associated with greater cumulative inflammatory burden (disease duration)^1^ and radiographic damage^5^. However, conclusive evidence of a causal role for inflammation through demonstration of reduction of CV risk with treatment of inflammation remains elusive. Historical incidence data^6–8^ has not shown the reductions in CVD expected with the advent of Biologic Disease-Modifying Antirheumatic Drugs (bDMARD). While cohort and registry data and uncontrolled studies indicates positive treatment effects conclusions must be tempered by constraints of the study design particularly selection bias^9^. Evidence from randomized controlled trials (RCT) is limited. Pooled analysis of secondary CV outcome data from RCTs in RA suggested a modest non-significant 15 percent reduction in CV events with TNF inhibition^9^. While much larger RCTs of canakinumab^10^ and cochicine^11^ have been found to reduce CV events by a similar order of magnitude in non-RA subjects these results cannot confidently be extrapolated to RA where these drugs have no efficacy. The low rate of clinical events in RA means that placebo controlled RCTs would have to be very large and of long duration. This has led many researchers to use pre-clinical CV biomarkers such as arterial stiffness and endothelial function (EF). Evaluating aortic stiffness one RCT found a better outcome in the placebo group over tocilizumab raising concerns regarding adverse effects of IL-6 inhibition upon lipids^12^ while another study found no difference between etanercept and placebo^13^.

## Objectives

This study evaluated the contribution of inflammation to CV risk in anti-cyclic citrullinated protein autoantibody (ACPA) positive RA by studying the effect of the TNF inhibitor adalimumab upon EF and arterial stiffness. The secondary aim of the study was to explore the relative contribution of RA disease-specific factors (disease stage, ACPA-titer and HLA-DR4 Shared Epitope (SE) status), smoking, blood pressure and lipid profiles upon these biomarkers.

## Methods

This single-site placebo controlled RCT (Clinical Trial Registry number: ACTRN12611000972921, Hunter New England Research Ethics Committee reference number 11/11/16/3.03) was funded by an Investigator-initiated grant from Abbvie and conducted in compliance with ethical standards outlined in the Declaration of Helsinki. The lead investigator obtained written informed consent from participants witnessed by a research assistant. Patients over 18 years of age with anti-CCP antibody (ACPA) positive RA^14^ and moderate to high disease activity were recruited from hospital outpatient clinics and consulting rooms between 27^th^ November 2012 and 21^st^ December 2015 across the city of Newcastle, Australia.

### Schedule of Assessments

Participants underwent two baseline assessments 2-7 days apart prior to being randomized 1:1 to receive treatment with adalimumab 40mg or placebo 2^nd^-weekly subcutaneous injections on a background of usual care. Randomization was performed by electronic random allocator in blocks of 20 prior to shipment of pre-filled syringes in sequentially numbered containers. The randomization code was held by the clinical trial pharmacist on site. Usual care was delivered by the treating clinician and could include conventional DMARDs, corticosteroids, NSAIDs and opiate analgesics. Participants then proceeded to assessments at 1, 4, 12 and 24 weeks of therapy during which time assessors and participants were blind to treatment. Following the Week 24 assessment the investigators and participants were un-blinded to treatment and this information conveyed to the treating clinician to inform subsequent treatment which could include bDMARDs. A single final open-label assessment was conducted between weeks 28 and 36 (hereafter referred to as Week 36).

### Assessments

Subjects were referred to the study with positive ACPA performed using commercially supplied second generation ACPA assays in government-accredited diagnostic laboratories. HLA typing was performed at Australian Red Cross Lifeblood (LABType SSO with Luminex System and One Lamba HLA Fusion software) to determine the dose (0,1 or2) of SE alleles^15^. Half-way through recruitment a protocol amendment was approved to measure serum ACPA and fasting lipid-profiles in participants at each time point in the study. These serial ACPA-assays were performed at University of Queensland Frazer Institute on patient serum with the QUANTA Lite CCP 3.1 IgG/IgA ELISA kit (Inova Diagnostics, Inc., San Diego, CA, USA) according to the manufacturer’s recommendations. Semi-quantitative anti-CCP concentrations were determined using a standard curve and the results are displayed as Units. Fasting lipid profiles were performed in the clinical laboratory at John Hunter Hospital. At each visit smoking status was recorded along with pulse, brachial artery blood pressure, height and weight. RA disease activity was evaluated by the principal investigator before proceeded to another room to undergo vascular assessments after resting supine for 5 minutes.

### Outcomes

#### 1. Endothelial Function (EF) – Primary Outcome

A trained research assistant assessed EF using the EndoPAT system^16^ to calculate Reactive Hyperemic Index (RHI) blind to treatment status and articular assessments. This automated system measures the increase in left index finger pulse amplitude following a 5-minute occlusion of blood flow with a pressure cuff inflated to 200mmHg. RHI is the ratio of post to pre-occlusion pulse amplitude adjusting for changes measured in the non-occluded right index finger. Greater values indicate better endothelial function. The effects of vascular occlusion can last 24 hours therefore a single assessment was performed at each time point.

#### 2. Arterial Stiffness – Secondary Outcome

The SphygmoCOR-XCEL system^17^ was used to perform automated measurements of central arterial pressures based upon right brachial artery pressure. Arterial stiffness was measured as Carotid-Femoral Pulse Wave Velocity (CF-PWV) calculated as the distance (meters) divided by the time delay (seconds) between the carotid and femoral pulses measured with a hand-held transducer (carotid) and femoral blood pressure cuff. Greater values indicate greater arterial stiffness and risk of CV events. Two assessments were performed at each encounter by the principal investigator blind to treatment.

#### 3. DAS28-CRP – Secondary Outcome

RA clinical disease activity was evaluated by 28 tender and swollen joint counts, patient global score of severity and CRP to calculate DAS28 scores^18^.

### Statistical Methods

Study size was calculated from published data for RHI in active RA (mean 2.02, standard error of the mean 0.07) versus inactive RA (mean 2.60, standard error of the mean 0.11)^4^. Assuming that inflammation was abrogated in all subjects treated with adalimumab it was calculated that a study with 13 patients in each arm could detect a significant difference (α 0.05, power 1-β 0.80). A recruitment target was set for 60 participants with ACPA-positive RA (30 with Early RA ≤ 12 months duration and 30 with Established RA > 12 months duration) to be randomized 1:1 Active (adalimumab) : Placebo within each group. The week 4 assessment was selected as the primary endpoint when differences in inflammatory burden would be greatest between the two groups.

Intra-observer reproducibility of DAS28-CRP and RHI were calculated by comparing the two Baseline assessments and summarized as the ICC by using a 2-way mixed effects regression with a random subject effect and a fixed effect indicating the first or second measure. The responsiveness was summarized as the smallest detectable difference (SDD) calculated as the two standard deviations of the difference scores for paired assessments^19^. For CF-PWV these statistics were calculated from the two measurements obtained at the Baseline 2 assessment due to the large changes observed between Baseline 1 and 2 in previous unpublished work.

For each outcome, we reported the means of the observed values for each treatment arm for the overall sample, as well as by disease stage (early or established). We also estimated the differences between treatment arms using a linear mixed model with unstructured residual variance to address within subject correlation and a random intercept for each participant. The first models regressed the outcome at each timepoint by treatment arm, time, an interaction term for treatment and time, baseline measure of the outcome, and stage of disease. In order to estimate differences between groups conditional on duration of treatment we fitted secondary models that also included a three-way interaction term for stage of disease, time and treatment, as well as all lower order interactions. Additionally, in accordance with recommendations modelling CF-PWV was adjusted for changes in central mean arterial blood pressure^20^.

Following this, we undertook analysis of area under the curve (AUC) (calculated by the trapezoidal rule) for each of DAS28-CRP, RHI and CF-PWV. Linear models were fitted to estimate the difference between the treatment groups in mean AUC for each outcome (univariate model). Additional multivariate models included explanatory variables of treatment group, baseline smoking status (Never / ex / current) and an interaction term between smoking and treatment (multivariate model 1) and treatment arm, SE status (0, 1 or 2 alleles) and an interaction term between SE status and treatment (multivariate model 2). No adjustment for multiple comparisons was made. Trends in ACPA titres and lipid profiles during the study period were evaluated for comparison with vascular biomarker assessments.

One participant was missing all baseline and post-baseline CF-PWV measures and was excluded from the main CF-PWV analyses. All other participants had complete outcome data for their second baseline measurement. Some, however, were missing post-baseline outcome measures and these were imputed with the mean of all of their non-missing measures from the first baseline measure to Week 36. All analysis was undertaken using R version 4.3.1.

## RESULTS

Sixty subjects (30 Early and 30 Established RA) were recruited from February 2012 to February 2016 (Figure 1). In the Early RA group, one patient in the placebo arm had a single baseline assessment. After week 12 six subjects (1 placebo, 5 active) became eligible for subsidized bDMARD therapy and were subsequently assessed open label. Two subjects withdrew from the study after week 24. In the Established RA group 3 subjects failed to return for follow-up assessments, one withdrew from the study after week 12 and one withdrew from the study upon diagnosis with lung cancer and was excluded from the analysis. One subject in the placebo arm with Established RA could not have assessments of CF-PWV due to obesity. After week 12 five subjects (2 placebo, 3 active) became eligible for government subsidized bDMARD therapy and underwent open-label assessment at weeks 24 and 36.

**Figure 1:**
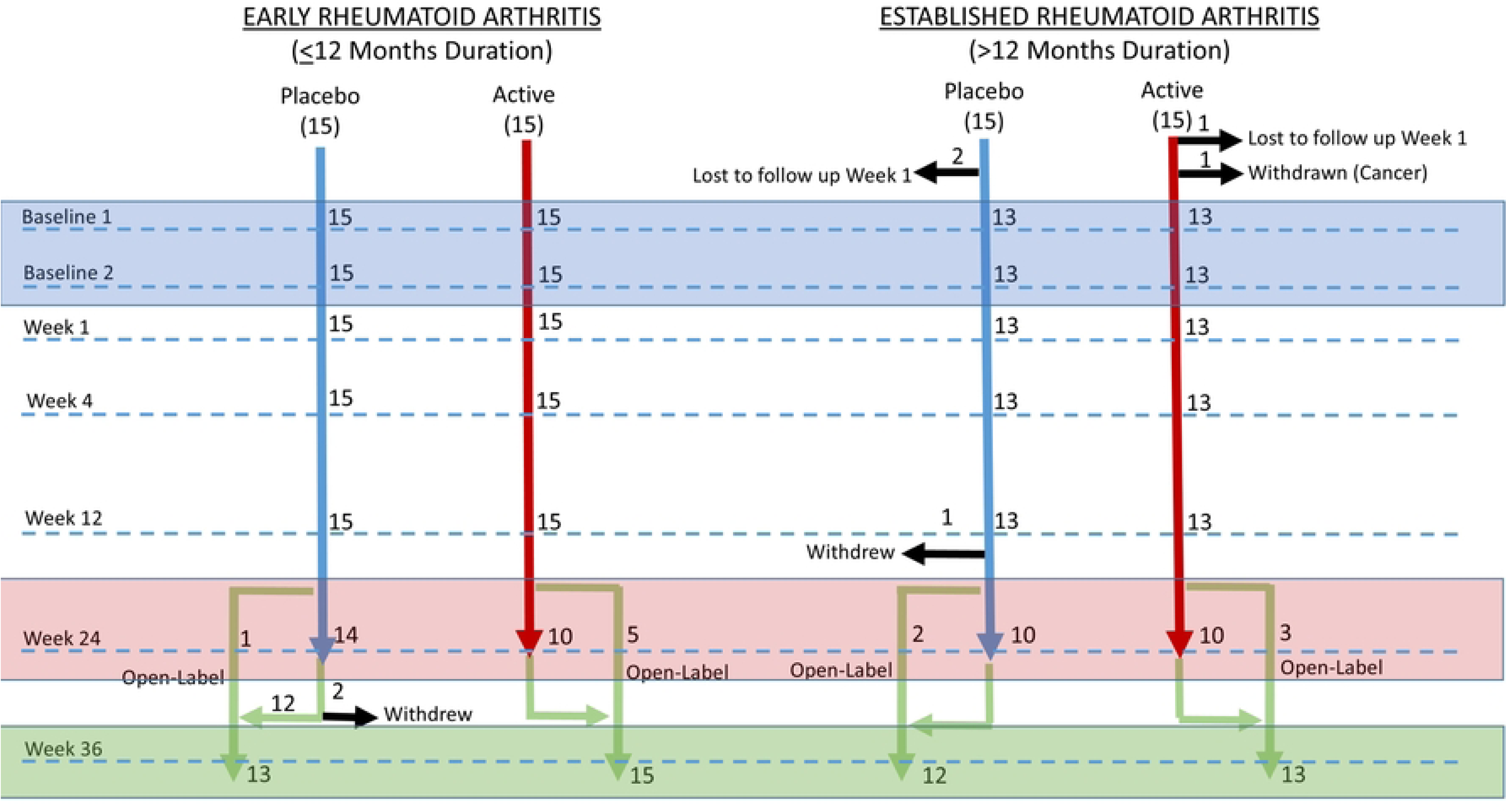
Study Flow Diagram.

Active and Placebo groups were similar at baseline (Table 1) although the placebo group had greater CF-PWV (second baseline 12.5 versus 11.8, p 0.178). The Active treatment group had more current smokers than placebo (35% versus 21%) but the two groups had similar numbers of “Ever Smokers” (82%). The groups were similar regarding ACPA titer and SE status. Reproducibility (ICC) was 0.732 for DAS28-CRP, 0.512 for RHI and 0.973 for CF-PWV. SDD was 1.89 units for DAS28-CRP, 0.96 units for RHI and 0.84 m/sec for CF-PWV.

**Table 1:** Descriptive Statistics at Baseline - Mean (Standard Deviation - σ) for continuous measures. Number (n) and percentage (in brackets) for categorical data.

| Characteristic |  | Treatment Group |  |
| --- | --- | --- | --- |
|  |  | Adalimumab (n=28) | Placebo (n=28) |
| Age (years) |  | 55.1 (12.3) | 57.8 (10.0) |
| Sex (Proportion Female) |  | 16/28 (57%) | 15/28 (54%) |
| Weight (kg) |  | 82.2 (17.2) | 80.6 (17.5) |
| Height (cm) |  | 168.1 (9.3) | 167.8 (8.2) |
| <u>Rheumatoid Arthritis Disease Characteristics</u> |  |  |  |
| Disease Duration (years) |  | 5.9 (8.6) | 5.4 (8.2) |
| Disease Group: | Early ($\leq 12$ months) | 15 (54%) | 15 (54%) |
| | Established ($>12$ months) | 13 (46%) | 13 (46%) |
| Rheumatoid Factor Titre (U/mL) |  | 345.7 (678.7) | 251.7 (304.3) |
| Anti-CCP Antibody Category (U/mL) | $\leq 30$ | 5 (18%) | 2 (7%) |
|  | 30-50 | 2 (7%) | 2 (7%) |
|  | 51-100 | 5 (18%) | 3 (11%) |
|  | 101-150 | 2 (7%) | 4 (14%) |
|  | 151-200 | 5 (18%) | 4 (14%) |
| | $>200$ | 9 (32%) | 13 (46%) |
| HLA-DR4 (Shared Epitope) Status | Negative | 4 (14%) | 3 (11%) |
|  | 1 Shared Epitope Allele | 12 (43%) | 13 (46%) |
|  | 2 Shared Epitope Alleles | 12 (43%) | 12 (43%) |
| Methotrexate: Proportion taking methotrexate |  | 24/28 (86%) | 25/28 (89%) |
| <u>Rheumatoid Arthritis Disease Activity</u> |  |  |  |
| 28 Tender Joint Count |  | 14.5 (10.2) | 14.6 (10.7) |
| 28 Swollen Joint Count |  | 9.4 (4.5) | 9.0 (4.3) |
| Entry CRP level (mg/L) |  | 33.8 (41.6) | 30.6 (33.3) |
| Entry ESR (mm/hour) |  | 26.5 (19.7) | 35.1 (25.9) |
| Physician Global Assessment (/100) |  | 52.4 (23.0) | 53.4 (20.9) |
| Patient Global Assessment (/100) |  | 52.7 (26.7) | 52.2 (21.5) |
| Simplified Disease Activity Index (SDAI) |  | 36.4 (17.0) | 36.6 (15.1) |
| DAS28-ESR |  | 5.6 (1.5) | 5.7 (1.5) |
| DAS28-CRP |  | 5.3 (1.3) | 5.3 (1.3) |
| 3-Variable DAS28-ESR |  | 5.4 (1.4) | 5.5 (1.4) |
| 3-Variable DAS28-CRP |  | 5.1 (1.2) | 5.1 (1.3) |
| <u>Conventional CV Risk Factors (excluding BP)</u> |  |  |  |
| Lipid Profile |  |  |  |
| High Density Lipoprotein (mmol/L) |  | 1.5 (0.6) | 1.2 (0.3) |
| Low Density Lipoprotein (mmol/L) |  | 3.2 (0.8) | 2.8 (0.8) |
| Triglyceride (mmol/L) |  | 1.5 (0.7) | 1.7 (0.9) |
| Smoking Status | Current smoker | 9 (32%) | 5 (18%) |
|  | Ex-smoker | 11 (39%) | 15 (54%) |
|  | Never smoked | 8 (29%) | 8 (29%) |
| <u>Cardiovascular Assessments</u> |  |  |  |
| Heart Rate (beats per minute) |  | 69.9 (12.5) | 67.8 (11.1) |
| Peripheral Systolic Blood Pressure (mm Hg) |  | 133.0 (13.2) | 134.8 (13.7) |
| Peripheral Diastolic Blood Pressure (mm Hg) |  | 79.2 (8.0) | 79.1 (8.7) |
| Central Systolic Pressure (mm Hg) |  | 121.8 (12.8) | 122.9 (12.1) |
| Central Diastolic Pressure (mm Hg) |  | 80.1 (8.1) | 79.8 (9.0) |
| Aortic Pulse Pressure (mm Hg) |  | 42.1 (12.6) | 43.2 (9.5) |
| <u>Cardiovascular Outcome Assessments</u> |  |  |  |
| Reactive Hyperaemic Index |  | 2.0 (0.5) | 2.1 (0.5) |
| Central Augmentation Index (%) |  | 20.1 (8.9) | 19.7 (7.4) |
| Carotid-Femoral Pulse Wave Velocity (m/sec) |  | 11.9 (1.8) | 12.5 (1.7) |

Figure 2 shows the unadjusted mean values with 95% confidence intervals for Placebo and Active arms at each time point for DAS28-CRP, RHI and CF-PWV with baseline-adjusted p-values for treatment versus placebo indicated (* p<0.05, ** p<0.01, *** p<0.001). Table 2 shows baseline-adjusted between-group differences. DAS28-CRP was lower in the adalimumab treated participants with early RA at weeks 1 (difference -1.72, 95% CI -2.73 to -0.71), 4 (difference -1.81, 95% CI -2.82 to

**Figure 2:**
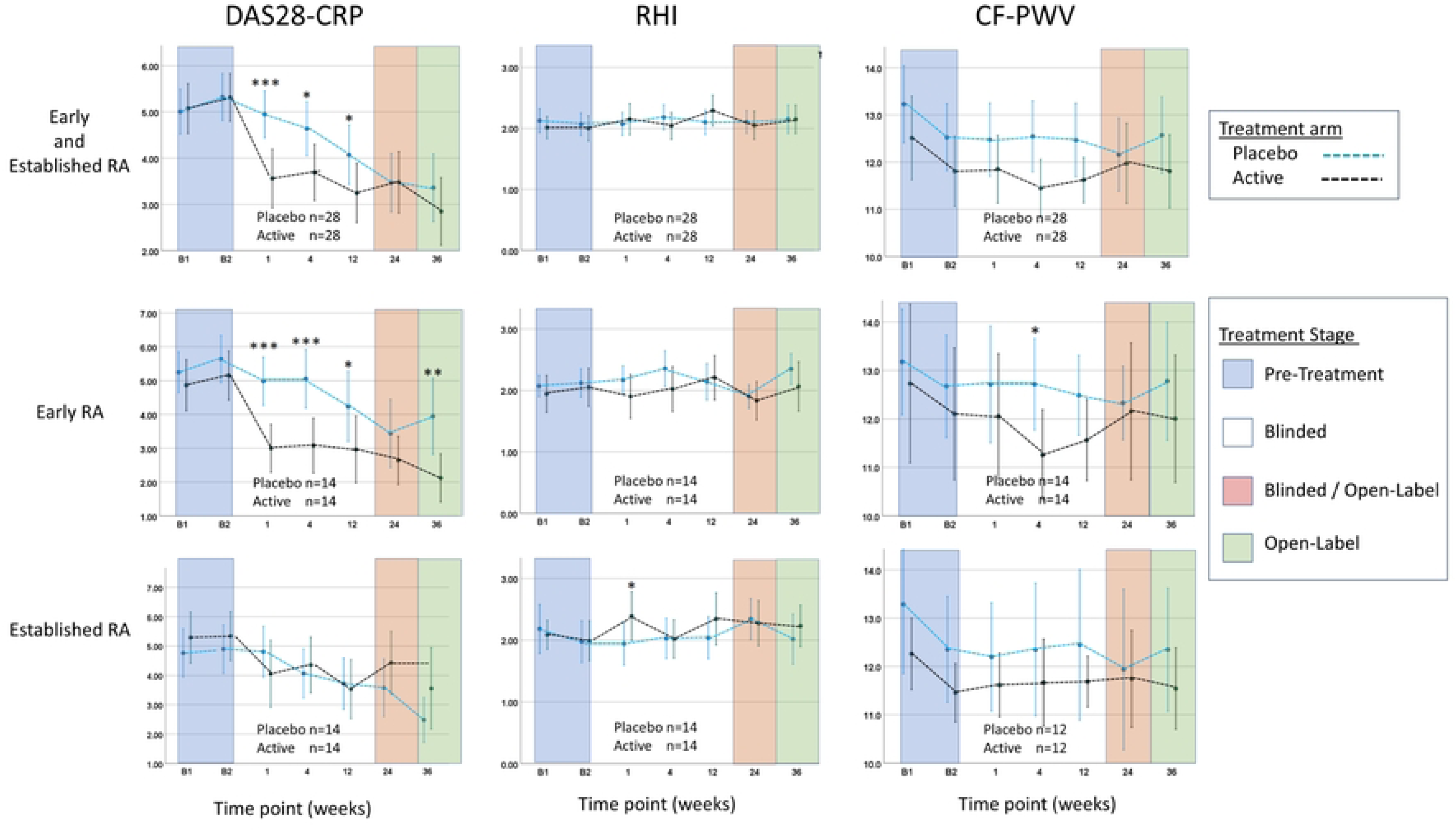
Treatment Outcomes. Graphs show unadjusted means.

**Table 2:**
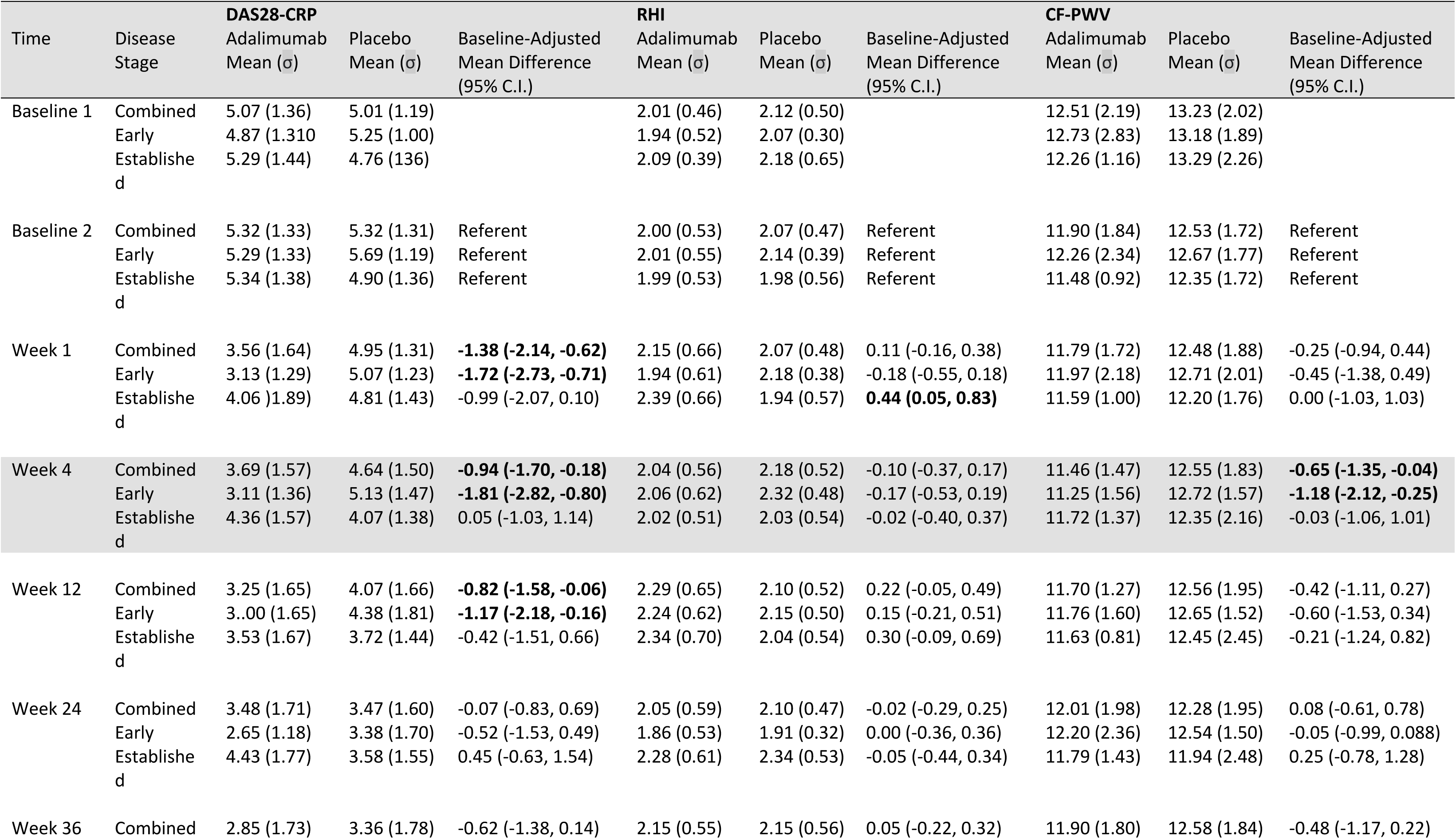
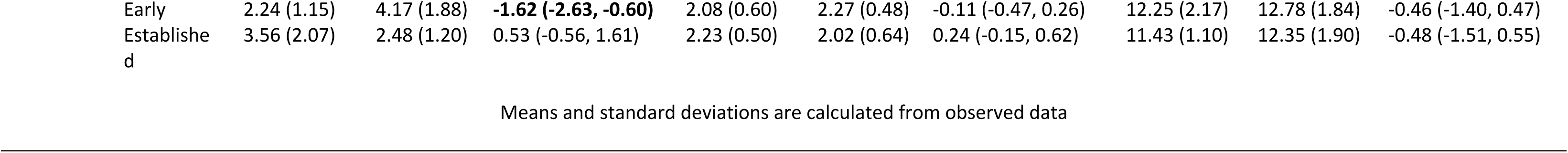
DAS28-CRP, Reactive Hyperemic Index (RHI) and Carotid-Femoral Pulse Wave Velocity (CF-PWV) means and standard deviations from the observed data (σ) for the two treatment groups with modelled baseline-adjusted differences and 95% confidence intervals. Significant results are indicated in bold type and the primary outcome in grey cells.

-0.80), 12 (difference -1.17, 95% CI -2.18 to -0.16) and 36 (difference -1.62, 95% CI -2.63 to -0.60) in Early RA while no treatment effect was seen in established RA. Better RHI was seen transiently in adalimumab subjects with established RA at week 1 (difference 0.44, 95% CI 0.05 to 0.83). CF-PWV saw a transient treatment effect in favour of adalimumab at week 4 in Early RA subjects (difference - 1.18 m/sec 95% CI -2.12 to -0.25). The greatest change in CF-PWV occurred between the two baseline assessments with an overall 0.74 m/sec reduction (p 0.052) occurring prior to treatment. Similar reductions were seen for peripheral blood pressures (systolic -5.7 mmHg, p 0.085 and diastolic -3.2 mmHg, p 0.034) and central blood pressure (systolic -4.9 mmHg p 0.053, diastolic -2.3 mmHg p 0.043 and mean -3.7 mmHg p 0.044).

Area under the curve analyses (Table 3) suggested that subjects who had never smoked had a trend to significant treatment effect for DAS28-CRP (difference -38.43, 95% confidence interval -83.94 to 7.08) and a significant treatment effect upon CF-PWV (difference -61.03, 95% confidence interval - 117.04 to -5.02). This was not seen in current or ex-smokers. Subjects expressing two SE alleles had no significant responses for DAS28-CRP (4.68, 95% confidence interval -38.03 to 41.39) or CF-PWV (13.48, 95% confidence interval -31.55 to 58.50). However, amongst subjects with one SE allele there was a trend to significant treatment effect for DAS28-CRP (difference -39.91, 95% confidence interval -76.07 to -3.75) and a significant treatment for CF-PWV (difference -48.82, 95% confidence interval -93.75 to -3.89). The small number of SE negative subjects showed a trend to treatment effects for DAS28-CRP (difference -31.53, 95% confidence interval -100.34 to 37.28) and CF-PWV (difference -68.42, 95% confidence interval -152.67 to 15.84). There was no evidence that the treatment affected RHI with an overall estimated difference of only 0.84 (95% confidence interval - 6.91 to 8.58) between the treatment arms. Nor was there evidence of treatment effect within any of the smoking or SE allele subgroups. Serial ACPA titer and lipid profiles did not significantly change over time in either treatment arm.

**Table 3:** Area Under the Curve - Estimated differences (with 95% confidence intervals) for the two treatment groups (Adalimumab versus Placebo n=28 for each group) for the DAS28-CRP, RHI and CF-PWV following multiple imputation of missing data.

| Model | Level | Number<br>(Active / Placebo) | **DAS28-CRP**<br>Estimated Difference<br>(95% C.I.) | p-value | **RHI**<br>Estimated Difference<br>(95% C.I.) | p-value | **CF-PWV**<br>Estimated Difference<br>(95% C.I.) | p-value |
| --- | --- | --- | --- | --- | --- | --- | --- | --- |
| Univariate |  | 28 / 28 | -19.85 (-45.08 to 5.37) | 0.120 | 0.84 (-6.91 to 8.58) | 0.829 | -24.67 (-55.16 to 5.81) | 0.111 |
| Multivariate (Model 1) |  |  |  |  |  |  |  |  |
| Smoking Status | Current | 9 / 5 | -27.79 (-78.60 to 23.03) | 0.277 | 3.47 (-13.07 to 20.02) | 0.675 | 25.77 (-36.88 to 88.43) | 0.413 |
|  | Ex | 11 / 15 | -10.28 (-46.57 to 26.01) | 0.572 | -2.96 (-14.74 to 8.83) | 0.617 | -22.12 (-67.04 to 22.79) | 0.327 |
|  | Never | 8 / 8 | -38.43 (-83.94 to 7.08) | 0.096 | 6.18 (-8.63 to 20.99) | 0.406 | **-61.03 (-117.04 to -5.02)** | **0.033** |
| Multivariate (Model 2) |  |  |  |  |  |  |  |  |
| Shared Epitope Status | 2 alleles | 12 / 12 | 4.68 (-32.03 to 41.39) | 0.799 | 0.88 (-11.23 to 12.99) | 0.885 | 13.48 (-31.55 to 58.50) | 0.550 |
|  | 1 allele | 12 / 13 | **-39.91 (-76.07 to -3.75)** | **0.031** | 3.26 (-8.65 to 15.16) | 0.585 | **-48.82 (-93.75 to -3.89)** | **0.034** |
|  | 0 alleles | 4 / 3 | -31.53 (-100.34 to 37.28) | 0.362 | -9.16 (-31.79 to 13.48) | 0.420 | -68.42 (-152.67 to 15.84) | 0.109 |

## Discussion

This is the first double-blind placebo-controlled RCT with the primary objective of evaluating the effect of TNF inhibition using adalimumab upon biomarkers of the risk of CVD in ACPA-positive RA that has been seen through to completion. There were striking treatment effects upon arthritis disease activity (DAS28-CRP) with adalimumab particularly in Early RA while there were only marginal and transient effects upon endothelial function (RHI) in Established RA at week 1 and upon CF-PWV at week 4 in Early RA. Additionally, the AUC analysis suggested that better DAS28-CRP and CF-PWV responses were seen in non-smokers and subjects with lower SE dose.

Our DAS28-CRP treatment responses support the existing evidence that better outcomes are generally seen in early RA^21^, non-smokers^22^ and in general with SE negative RA^23,24^ although better treatment responses are seen specifically with abatacept in SE positive RA^25^. The better CF-PWV response in subjects with lower SE dose and non-smokers mirror the arthritis treatment responses and is consistent with the evidence that SE-positive RA is associated with greater risk of CVD^5,26^. The similarities in DAS-CRP and CF-PWV responses appear to support the hypothesis that SE status, inflammation, disease duration and smoking status determine the severity of both the inflammatory arthritis and CVD. However, any conclusions regarding potential mechanisms must be informed by an understanding of the limitations of the CV biomarkers and the emerging complexity of disease mechanisms responsible for excess CVD in RA.

### 1. Limitations of Outcome Assessments

#### a. Endothelial Function

RHI has been found to be predictive of CV events in very large high-risk non-RA cohorts^27^. A previous uncontrolled study^4^ suggested that RHI would be a useful outcome assessment in RA and the reproducibility of RHI in our study was similar to that previously reported^28^ suggesting that our study was adequately powered. While our study appeared to confirm a beneficial treatment effect this was marginal and transient while there was a clear and sustained reduction in inflammatory burden as measured by DAS28-CRP. The RHI result therefore more likely represents a type 1 statistical error.

EF is a complex phenomenon that has an unclear relationship with inflammation in RA. Some studies have found poor EF in RA compared to healthy controls^29^ while others have not^30^. Some studies reported poorer EF in active versus inactive RA^4^ while others have not^31^. One large study paradoxically found the best EF in subjects with greatest disease activity^29^ contradicting strong epidemiological evidence of association between high disease activity and CV events^3^. The variability of these results may reflect differences in study populations or in methods for evaluating EF. Our findings suggest that RHI does not correlate with inflammatory burden in RA and is not a useful assessment to evaluate the contribution of inflammation to excess CVD in RA.

#### b. Carotid-Femoral Pulse Wave Velocity

CF-PWV is a robust predictor of CV events in non-RA subjects. A 1 m/sec increase in CF-PWV is associated with 14% elevation in CV events^32^. CF-PWV has consistently been found to be around 1 m/sec greater in RA compared to healthy control subjects^29,33,34^. CF-PWV is a direct measure of aortic and large vessel stiffness^35^. Increases in CF-PWV reflect structural changes such as collagen proliferation, elastin fragmentation and calcification that occur as a result of cumulative micro-trauma with each cardiac cycle^36^. As vessel walls stiffen, they become less able to accommodate biomechanical stresses incurring greater micro-trauma resulting in a positive feed-back loop so that CF-PWV increases exponentially through life^20,37^. However, CF-PWV is also influenced by other factors. Simply increasing the intra-luminal blood pressure distends blood vessels increasing wall tension moving the vascular wall biomechanical responses into a less accommodating (stiffer) part of the stress-strain response curve following mathematical relationships first described a century ago^38^. The arterial pulse pressure wave is then propagated at greater velocity without any change in the structural properties of the vessel wall. This effect was seen very clearly in our study where large reductions in CF-PWV occurred between the two assessments <u>prior</u> to treatment concurrently with reductions in blood pressure due to reductions in “white coat hypertension”. Our data suggests that two assessments of CF-PWV should be performed on separate days, the first simply to familiarize subjects with the procedure so that the second and subsequent measurements used for analysis once “white coat hypertension” has settled. Such physiological mechanisms may account for apparent reductions in arterial stiffness seen in uncontrolled trials^33,39–41^, and those seen in both arms of non-randomized^42^ and randomized^43^ controlled trials where only one baseline assessment was performed prior to treatment. The largest RCT, evaluating CF-PWV as a secondary outcome, found transient reductions in CF-PWV in the placebo arm of the study more consistent with physiological than structural mechanisms^12^. It is now recommend that CF-PWV be adjusted for central mean arterial pressure^20^. While we statistically adjusted for changes in mean arterial blood pressure using linear modelling, the transient positive treatment effects we observed for CF-PWV in the adalimumab treated group are more consistent with changes in blood pressure due to reduction in pain. As blood pressure is not linearly related to CF-PWV linear modelling may not have sufficiently accounted for this effect. In our study, this mechanism would better account for the significant treatment effects for CF-PWV were seen in subjects with better DAS-CRP responses in subjects with early disease, lower SE dose and non-smokers as they had greater reduction in arthritis disease activity and in pain.

Despite these limitations CF-PWV may yet prove to be a valuable biomarker for RA research. CF-PWV reflects cumulative arterial damage and is somewhat analogous to radiographic Sharp scores^19^. Viewed this way it becomes apparent that clinical trials should be designed to detect differences in rates of progression. An added layer of complexity is that CF-PWV normally increases in mid-life at a rate of 0.1 m/sec/year^44^. While the longitudinal trajectories of CF-PWV have not been studied in RA it is known that subjects with RA of 10 years duration have mean CF-PWV 1.0 m/sec greater than healthy controls^29^. Assuming that subjects have normal CF-PWV prior to onset of RA, we may infer that the trajectories of RA and Healthy populations diverge at a rate of 0.1 m/sec/year. Assuming that that this divergence is entirely due to inflammation, that treatment completely abrogates inflammation and that this brings the rate of increase in CF-PWV in line with that of the general population we might expect the trajectories of active and placebo arms in a controlled trial to also diverge at a rate of 0.1 m/sec/year. Based upon the performance metrics of CF-PWV in our study 161 subjects would be required in each arm of a 1:1 placebo controlled RCT to detect a difference of 1.0 m/sec at 10 years. CF-PWV may be better suited for use in cross-sectional observational studies where it serves as a summary measure of a range of contributors to arterial stiffness accumulated over a lifetime. With the emerging understanding of the mechanistic complexity of excess CV in RA this could be the strength of CF-PWV assessments.

### 2. Complexity of Disease Mechanisms

The emerging complexity of the mechanisms of excess CVD in RA presents additional challenges to research in this area. Studies in this field have generally included ACPA-negative RA where disease mechanisms may differ. It is also emerging that inflammatory burden alone may not fully account for excess CVD in RA. The advent of drugs that effectively abrogate inflammation has not seen the expected 30-50% reductions if inflammation was the principal cause of excess CVD^6,7^. Pooled data from three major TNF inhibitor RCTs^9^ suggested that TNF inhibitors might reduce CV events by a more modest 15% although this result was not statistically significant. Similarly, in non-RA subjects canakinumab^10^ and colchicine^11^ might reduce CV events by 10-15%. These figures suggest that inflammatory mechanisms might account for one third of the excess CVD in RA. This is broadly consistent with epidemiologic evidence that 49% of CV risk can be attributed to conventional risk factors while RA specific factors (inflammation and autoimmunity) account for another 20% leaving 30% unaccounted for^2^. It is unclear to what extent the elevation in CF-PWV is a result of inflammation. The 1 m/sec elevation of CF-PWV seen in RA^29^ would be associated with a 15% greater risk of CV events^32^ that may form part of that unexplained part of the CV risk.

One intriguing observation is that CV risk is elevated prior to the onset of inflammatory arthritis^45,46^. It is now understood that ACPA-positive RA arises from interaction between genetic (HLA-DR4 Shared Epitope) and environmental factors. A pre-clinical state of autoimmunity (ACPA positivity) and migratory arthralgia can exist for decades^47^. As the threshold of clinical RA is approached there is a broadening of the ACPA repertoire^48^ and increases in serum cytokines^49^. These processes might explain much of the pre-clinical increase in CV risk. However, it is notable that many of the modifiable risk factors for the development of RA are also risk factors for CVD hinting at the possibility of non-immunological common origins of the two diseases. One study^50^ has explored the hypothesis that RA and CVD have common biomechanical origins. The study suggested that, while autoimmunity lays the foundation for RA, articular stiffness then determines the severity of RA. Arterial stiffness correlates with articular stiffness and therefore “constitutional stiffness” is a common determinant of RA severity and CV risk.

This study demonstrated transient treatment effects of adalimumab upon two biomarkers of CVD in RA that are unlikely to represent true direct effects of inflammation. This study highlights the challenges of research in this area. A better-informed understanding of the pathophysiology of these assessments may identify identifies avenues for future research exploring the origins of excess CVD in RA.

## Data Availability

Data will be shared upon reasonable request to the corresponding author. Study data are the property of NSW Department of Health and will be released via secure transfer mechanisms in accordance with NSW Government Department of Health policy.

## Data Availability

Data will be available from request.

## Statement of ethics and consent

Informed written consent was obtained from all participants in this study. The study was conducted in compliance with ethical standards outlined in the Declaration of Helsinki

